# Elevated antiviral, myeloid and endothelial inflammatory markers in severe COVID-19

**DOI:** 10.1101/2020.10.08.20209411

**Authors:** Ryan S Thwaites, Ashley Sanchez Sevilla Uruchurtu, Matthew Siggins, Felicity Liew, Clark D Russell, Shona C Moore, Edwin Carter, Simon Abrams, Charlotte-Eve Short, Thilipan Thaventhiran, Emma Bergstrom, Zoe Gardener, Stephanie Ascough, Christopher Chiu, Annemarie B Docherty, David Hunt, Yanick J Crow, Tom Solomon, Graham P Taylor, Lance Turtle, Ewen M Harrison, Malcolm G Semple, J Kenneth Baillie, Peter JM Openshaw, on behalf of the ISARIC4C investigators

## Abstract

The mechanisms that underpin COVID-19 disease severity, and determine the outcome of infection, are only beginning to be unraveled. The host inflammatory response contributes to lung injury, but circulating mediators levels fall below those in classical ‘cytokine storms’. We analyzed serial plasma samples from 619 patients hospitalized with COVID-19 recruited through the prospective multicenter ISARIC clinical characterization protocol U.K. study and 39 milder community cases not requiring hospitalization. Elevated levels of numerous mediators including angiopoietin-2, CXCL10, and GM-CSF were seen at recruitment in patients who later died. Markers of endothelial injury (angiopoietin-2 and von-Willebrand factor A2) were detected early in some patients, while inflammatory cytokines and markers of lung injury persisted for several weeks in fatal COVID-19 despite decreasing antiviral cytokine levels. Overall, markers of myeloid or endothelial cell activation were associated with severe, progressive, and fatal disease indicating a central role for innate immune activation and vascular inflammation in COVID-19.

## Main text

Fatal COVID-19 is associated with acute respiratory distress syndrome and raised systemic inflammatory markers including IL-6 and C-reactive protein, often accompanied by neutrophilia and lymphopenia ^1^. The beneficial effect of corticosteroid treatment in severe disease highlights the role of steroid-responsive inflammation in pathogenesis ^2, 3^, and post-mortem studies report pulmonary vessel vasculitis (most commonly myeloid cells) and microthrombosis in fatal COVID-19 ^4, 5, 6, 7^. The virus-induced inflammatory state has laboratory features that resemble secondary haemophagocytic lymphohistiocytosis (sHLH) ^8, 9, 10^ but the exact pattern and severity of inflammatory responses has been only partially characterized. Levels of some inflammatory mediators, including IL-6, are elevated in COVID-19, but are typically ten times lower than those reported in acute respiratory distress syndrome (ARDS) and sepsis ^11, 12, 13^, suggesting that other factors may play a major role in COVID-19 severity. Host genetic factors may also influence disease severity, with polymorphisms in several regions, including the interferon pathway genes *IFNAR2* and *OAS1/2/3* recently associated with enhanced disease severity ^14^. Identification of such genetic of inflammatory factors may define a ‘treatable trait’ ^15^, allowing both stratification of patients likely to benefit from therapies such as dexamethasone and targeted biological anti-cytokine therapies, and design of novel therapeutics targeting causative pathways.

Early clinical studies of COVID-19 identified elevated neutrophil counts and lymphopenia in peripheral blood ^1, 16^, predominantly seen in late-stage disease and of limited prognostic value. Peripheral blood neutrophilia is also seen in other severe respiratory viral ^17^ and bacterial ^18^ infections, suggesting that this is not a unique feature of COVID-19. Elevated levels of D-dimer, a product of fibrin-degradation associated with thrombosis and inflammation, have also been observed in COVID-19 ^16^, consistent with systemic inflammation and the high frequency of macrovascular thrombotic complications in severe cases ^7, 19^. Post-mortem studies show that thromboses and microthrombi within pulmonary vessels are common in fatal COVID-19 and are associated with endothelial responses distinct from those that occur during fatal influenza A virus infection ^4, 5, 7, 20^. However, the thrombotic aspects of life-threatening COVID-19, and the interaction of this process with cytokine release have hitherto been described in relatively small groups of cases, from single-center studies, or with a narrow range of disease severities.

Within the ISARIC4C study we obtained clinical data and 1,047 plasma samples from 619 hospitalized patients with COVID-19 ^21, 22^. Given the large number of cases, patients from the ISARIC4C database could be stratified into five levels of severity according to their peak illness, in line with the World Health Organization COVID-19 ordinal scale ^23^ (Supplementary Table 1): (1) no oxygen requirement (Severity 3, n=169); (2) patients requiring oxygen by face mask (Severity 4, n=143); (3) patients requiring high-flow nasal cannulae, a continuous positive airway pressure mask or other non-invasive ventilation (Severity 5 n=99); (4) patients requiring invasive mechanical ventilation (Severity 6/7, n=113); and (5) fatal COVID-19 (Severity 8, n=95). The median duration of symptoms prior to study enrollment was similar in all groups: Severity 3, 7 days; Severity 4, 9 days; Severity 5, 11 days; Severity 6/7, 11 days; and Severity 8, 8 days. Some differences in routinely performed clinical hematology and biochemistry measures were evident between clinical outcome groups at the time of study enrolment: Lymphopenia was evident in groups 6/7 and 8, relative to 3, alongside neutrophilia in 6/7 and 8 relative to 3 and 4 (Supplementary Fig. 1a and 1b, respectively). No differences between groups were observed in ferritin levels, whilst LDH was elevated in groups 5, 6/7, and 8 relative to 3 (Supplementary Fig. 1c and 1d, respectively). Procalcitonin levels were elevated in group 8 relative to 3 and 4, and in group 6/7 relative to 4 (Supplementary Fig. 1e). Partial HScores ^24^ were calculated (fever, cytopenia, ferritin, triglycerides, and AST) but the only significant difference between groups was between 6/7 and 4, indicating that sHLH is unlikely to be the predominant pathophysiological mechanism in life-threatening COVID-19 (Supplementary Fig. 1f). The ISARIC4C mortality scores ^25^ for these patients demonstrated an elevated risk of mortality, calculated from admission data, in those that would progress to fatal disease (group 8) relative to all other groups (Supplementary Fig. 1g), though there was considerable overlap between all groups. Together, these data indicated limited clinical or biochemical differences between patient outcome groups at the time of hospital admission.

We hypothesized that differences in the levels of plasma inflammatory mediators would reflect the nature and scale of immunopathology in COVID-19 and would associate with different disease outcomes. We therefore quantified 33 mediators in all available plasma samples using panels designed to study a broad range of mediators that could be broadly categorized as having roles in antiviral immunity, inflammation, or coagulation ^16, 19^. Analysis of plasma mediator levels at the time of enrolment distinguished 3 clusters of patients that were associated with distinct patterns of mediator levels (Fig. 1). The first of these clusters was enriched in patients from groups 6/7 and 8 and was associated with higher levels of CXCL10, GM-CSF, D-dimer, and vWF-A2. The second cluster contained a more diverse mixture of severities and had a more pronounced pattern of coagulation factor XIV and angiopoietin-2 containing mediator clusters, but lower levels of the CXCL10 containing mediator cluster. The third patient cluster had lower levels of the CXCL10, D-dimer, and coagulation factor XIV containing mediator clusters, but had a more varied pattern of other mediators including IL-6Rα, VEGF-D, and IL-4. Interestingly, this analysis did not indicate any obvious patterns of age, symptom duration (onset), or sex in these plasma mediator levels. This analysis shows that, at the time of enrolment, different COVID-19 outcome groups were already identifiable and associated with distinct patterns of inflammatory mediators and that markers such as D-dimer, EN-RAGE, CXCL10, and GM-CSF were particularly associated with enhanced disease severity. However, entry to the study was determined by hospitalization which will be influenced by predisposing factors; these factors may therefore not be evident in the data that we accumulate.

**Fig. 1.**
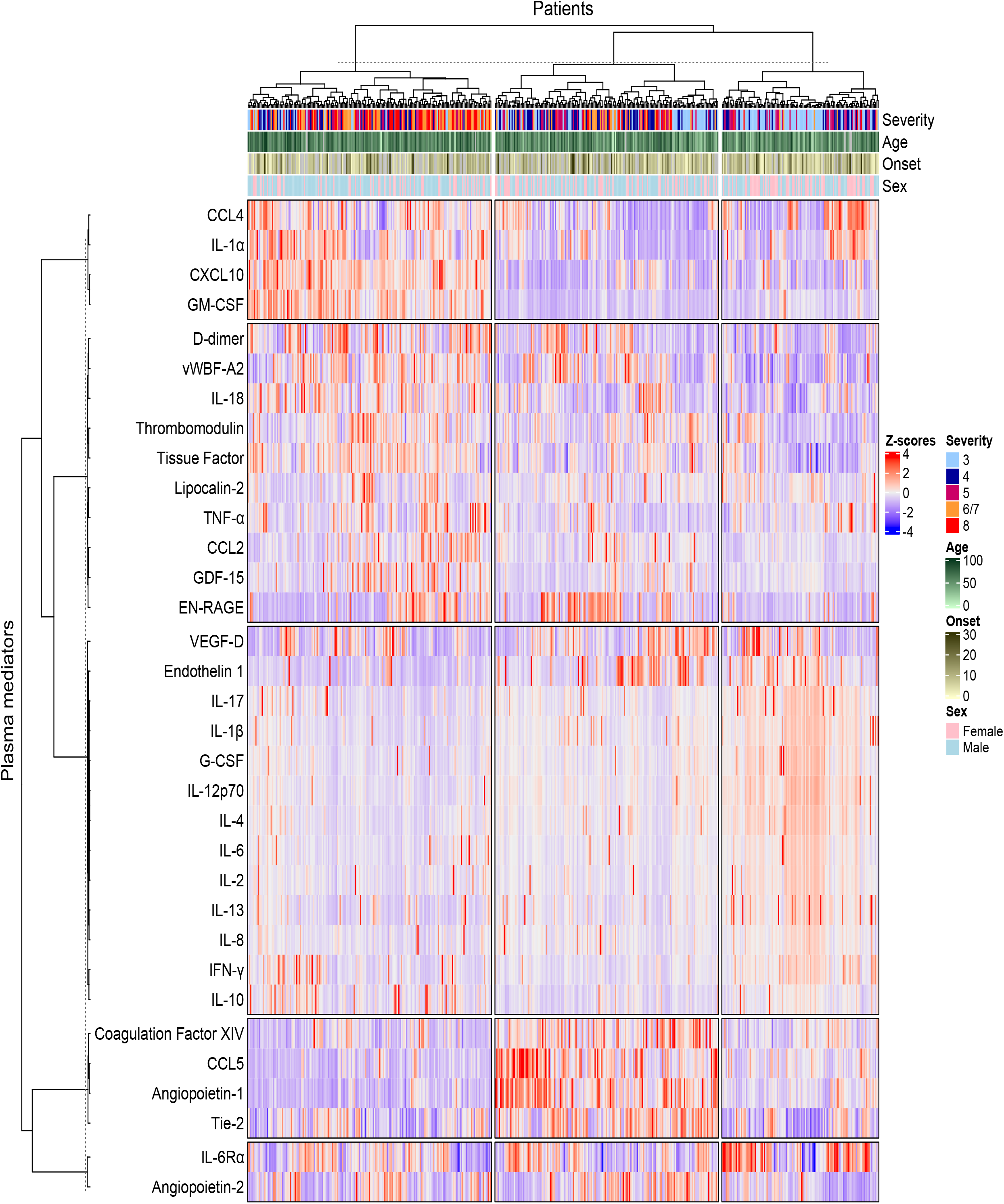
Plasma mediators at the time of study enrollment demonstrate a broad exaggerated immune response in patients hospitalized with COVID-19. Clustered heatmap of 33 immune mediators in plasma samples collected from patients hospitalized with COVID-19 at the time of study enrolment. Values for each mediator were scaled and rows and columns were split by K-means clustering. Each patients’ column is additionally annotated with data on disease outcome (“Severity”) as one of the following outcome groups: not requiring oxygen support (‘3’, n=128), requiring oxygen via a face mask (‘4’, n=103), requiring non-invasive ventilation or high-flow nasal canulae (‘5’, n=78), requiring invasive mechanical ventilation (‘6/7’, n=87) or fatal disease (‘8’, n=69). Columns are additionally annotated with patient age, sex and duration of illness at the time of sample collection (“Onset”).

To further explore the relationship between the mediator levels and severity we analyzed plasma from 15 healthy controls (7 males, median age 55, range 45-71) and 39 individuals recruited 7 days after a SARS-CoV-2 positive PCR test who did not require hospitalization (15 males, median age 43, range 27-62, termed group ‘1/2’ as per the WHO scale ^23^) and related these to hospitalized patients. At the time of enrollment, numerous differences were evident between hospitalized COVID-19 patients and the control groups, along with many differences across the clinical outcome groups in hospitalized patients (Fig. 2 and Supplementary Fig. 2). In contrast to other reports ^26^ we found no evident deficiency in IFN-α levels in those with severe disease (Fig. 2a). IFN-γ was elevated in hospitalized COVID-19 patients relative to both healthy controls (HC) and group 1/2 (Fig. 2b) and was elevated in the most severe outcome groups, relative to lower severity grades. The interferon-induced chemokine CXCL10 was also substantially elevated in all hospitalized COVID-19 cases relative to the control groups, with the most pronounced increases in groups 6/7 and 8 (Fig. 2c). These results are in contrast to the decreased ISG gene expression in peripheral blood samples from patients with severe COVID-19 ^26^, showing that the gene expression pattern from blood does not necessarily reflect the directly measured levels of gene product. We speculate that the abundance of IFN-γ and CXCL10 results from release from the site of disease rather than from circulating cells, though anti-IFN autoantibodies ^27^ and polymorphisms in IFN signaling ^14^ may influence this pathway in some patients.

**Fig. 2.**
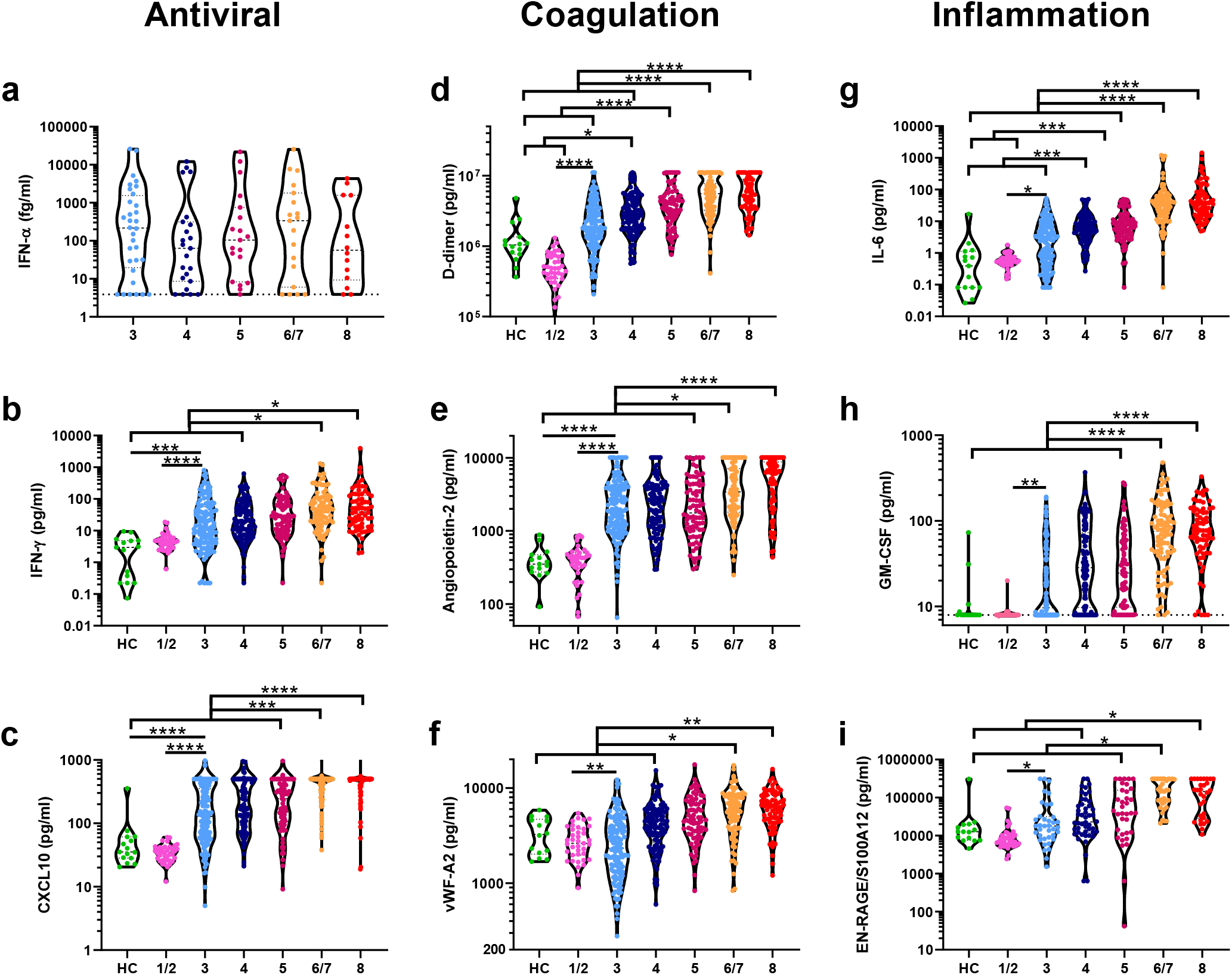
Antiviral, coagulation, and inflammation associated mediators distinguish severity groups early in disease. Plasma samples from the time of study enrolment were analyzed for levels of the antiviral cytokines a) IFN-α, b) IFN-γ, and c) the interferon-induced chemokine CXCL10 in healthy control (HC, n=15), patients with COVID-19 not requiring hospitalization (‘1/2’, n=39), and hospitalized patients with COVID-19 that would: not require oxygen support (‘3’, n=32-128), require an oxygen face mask (‘4’, n=23-103), require non-invasive ventilation or high-flow nasal cannulae (‘5’, n=19-78), require invasive mechanical ventilation (‘6/7’, n=19-87) or progress to fatal disease (‘8’, n=14-69). Mediators associated with coagulation and endothelial injury were also quantified in these plasma samples; d) D-dimer, e) Angiopoietin-2, and f) von-Willebrand factor A2 (vWF-A2). Similarly, mediators associated with inflammation were quantified: g) IL-6; h) GM-CSF; and i) EN-RAGE/S100A12. Data were analyzed for statistical significance using Kruskal-Wallis tests with Dunn’s tests for multiple comparisons between all groups. \**P*<0.05, \*\**P*<0.01, \*\*\**P*<0.001, \*\*\*\**P*<0.0001.

The fibrin degradation product D-dimer has been reported to be elevated in severe COVID-19 ^16^, implicating thrombosis in disease severity ^4, 5, 20^. In agreement with these reports, D-dimer was elevated in all hospitalized groups, but little difference was observed between the severity groups at the time of enrolment (Fig. 2d). Given reports of the association between COVID-19 mortality and pulmonary vasculitis ^4^, we hypothesized that endothelial injury may be a feature of COVID-19, potentially triggering coagulation and the thrombotic complications common in severe disease ^19, 28^. Indeed, levels of angiopoietin-2, a marker of endothelial injury, were elevated in all hospitalized patients relative to both control groups (Fig. 2e), with levels 5.6-fold higher in the mildest hospitalized patients (group 3, median=1983pg/ml) than HCs (median=352pg/ml). Angiopoetin-2 levels were also significantly elevated in groups 6/7 and 8 relative to all other hospitalized COVID-19 outcome groups (Fig. 2e). As both angiopoietin-2 and vWF-A2 can enter the blood plasma through exocytosis of endothelial cell Weibel-Palade bodies ^29^, we also quantified vWF-A2, which was similarly elevated in hospitalized COVID-19 patients (Fig. 2f). In line with these markers of endothelial injury and thrombosis, thrombomodulin, vWF-A2, and endothelin-1 were also elevated in COVID-19, predominantly in those most severe patient outcome groups (Supplementary Fig. 2). Elevations in these prothrombotic mediators were not counteracted by levels of the inhibitors angiopoietin-1 or soluble Tie2, which were not significantly different between the tested groups (Fig. S2). These results suggested that endothelial injury and coagulation are common features of patients hospitalized with COVID-19 and that these are most pronounced in severe and fatal COVID-19.

In line with other reports ^1, 12^, we found that IL-6 was also significantly elevated in most hospitalized groups relative to the controls (Fig. 2g), with a stepwise increase in levels with escalating severity. IL-6 levels in groups 6/7 and 8 were significantly elevated above all other groups (all *P*<0.0001, Fig. 2g). In agreement with the association of a strong inflammatory response with COVID-19 severity, GM-CSF was similarly elevated in all hospitalized groups, relative to controls and was most pronounced in the groups 6/7 and 8 (Fig. 2h). Numerous other inflammatory cytokines and chemokines showed similar results including TNF-α, IL-2, GDF-15, G-CSF, and VEGF-D (Supplementary Fig. 2). EN-RAGE/S100A12 has previously been characterized as a marker of respiratory damage in ARDS ^30^ and indeed was elevated in groups 6/7 and 8 relative to most others (Fig. 2i). The neutrophil chemokine IL-8 (CXCL8) was similarly elevated in severe disease, as was the neutrophil gelatinase associated lipocalin (LCN-2/NGAL) (Supplementary Fig. 2), in line with the reported association between blood neutrophilia and severity ^16^ also seen in this cohort (Supplementary Fig. 1b).

Other immunological mediators (IL-6Rα, IL-13, IL-17) were not significantly different between groups, indicating that only limited aspects of the immune repertoire were active in COVID-19. Interestingly, IL-4 levels were lower in the non-severe disease outcome groups (3, 4, and 5) relative to both the control groups and the severe disease groups 6/7 and 8 (Supplementary Fig. 2), indicating that suppression of the normal levels of type-2 cytokines may be associated with milder COVID-19 disease, and that this mechanism is lost in severe disease. Similarly, IL-12p70, commonly released by antigen presenting cells (APCs) ^31^, was decreased in all hospitalized cases relative to the HCs and group 1/2 (Figure S2), possibly owing to the trafficking of APCs to the site(s) of viral infection.

To determine the strength of the relationships between these individual plasma mediators we performed a hierarchical correlation matrix analysis of mediators from plasma samples collected at the time of study enrolment. This identified a strongly correlated cluster of inflammatory mediators including GM-CSF, CXCL10, vWF-A2, and IL-6 (Fig. 3a); increases in which were commonly associated with the most severe COVID-19 outcome groups. Given the strong association between age and COVID-19 severity ^22^, and reports of increased inflammatory responses in males relative to females with COVID-19 ^32^ we investigated the influence of these demographic factors on plasma mediators levels in hospitalized patients. As the major effect in our cluster analysis was severity (Fig. 1), we further stratified each of these severity groups by age (≥ or < 70 years of age) and sex, to better account for the influence of disease severity on plasma mediator levels. Following adjustment for multiple testing, no mediator was found to be statistically different between males and females within each outcome group (Supplementary Fig. 3). By contrast, several differences were evident between those aged ≥70 and <70 years, including elevated levels of D-dimer, CXCL10, and GM-CSF in those aged ≥70 years; IFN-γ levels were, by contrast, greater in younger patients within severity group 4 (Fig. 3b and Supplementary Fig. 3).

**Fig. 3.**
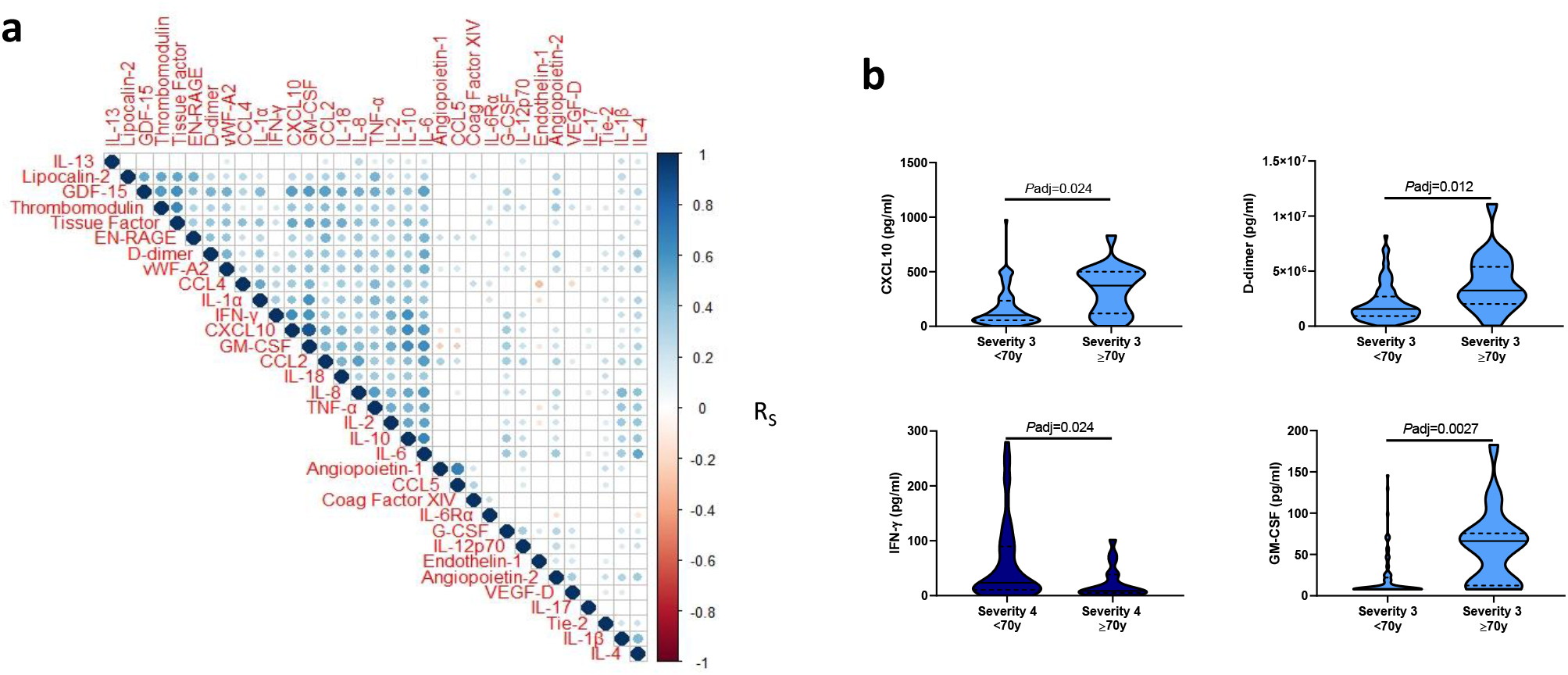
Plasma mediators in COVID-19 are coordinated around GM-CSF and influenced by age. a) Correlogram of the association between plasma mediator levels at the time of enrolment in all patients hospitalized with COVID-19 (n=465). b) Inflammatory mediator levels within an outcome group, stratified as those ≥ or < than 70 years of age. Data in panel a were analyzed using Spearman’s rank correlations with correction for multiple testing; significant correlations are denoted by a circle, the color of which denotes the Spearman’s R value. Data in panel b were analyzed using Mann-Whitney U tests with *P*-value adjustment for false discovery rate.

We next sought to determine the changes in levels of some key plasma mediators from the time of enrolment over the course of disease, by relating data to the patient reported duration of symptoms at the time of each sample collection, including consecutive samples collected from individual patients. This analysis indicated that many mediators were stable over the time-course of hospitalization, supporting the validity of using samples from the time of enrolment to study the immunologic basis of COVID-19. However, some mediators did change over time; for example, there was a gradual decrease in IFN-γ and CXCL10 over time in most groups (Supplementary Fig. 4), including group 8 (Fig. 4a and 4b, respectively). By contrast some other mediators remained elevated or appeared to increase over the duration of symptoms in group 8, including angiopoietin-2 and D-dimer (Fig. 4c and 4d, respectively). Similarly, the inflammatory mediators GM-CSF and EN-RAGE remained elevated or increased in group 8 in the latter stages of disease (Fig. 4e and 4f, respectively). Together, these results indicated that the most severe outcomes of COVID-19 disease were associated with persistent coagulation and inflammation, even as IFN levels declined.

**Fig. 4.**
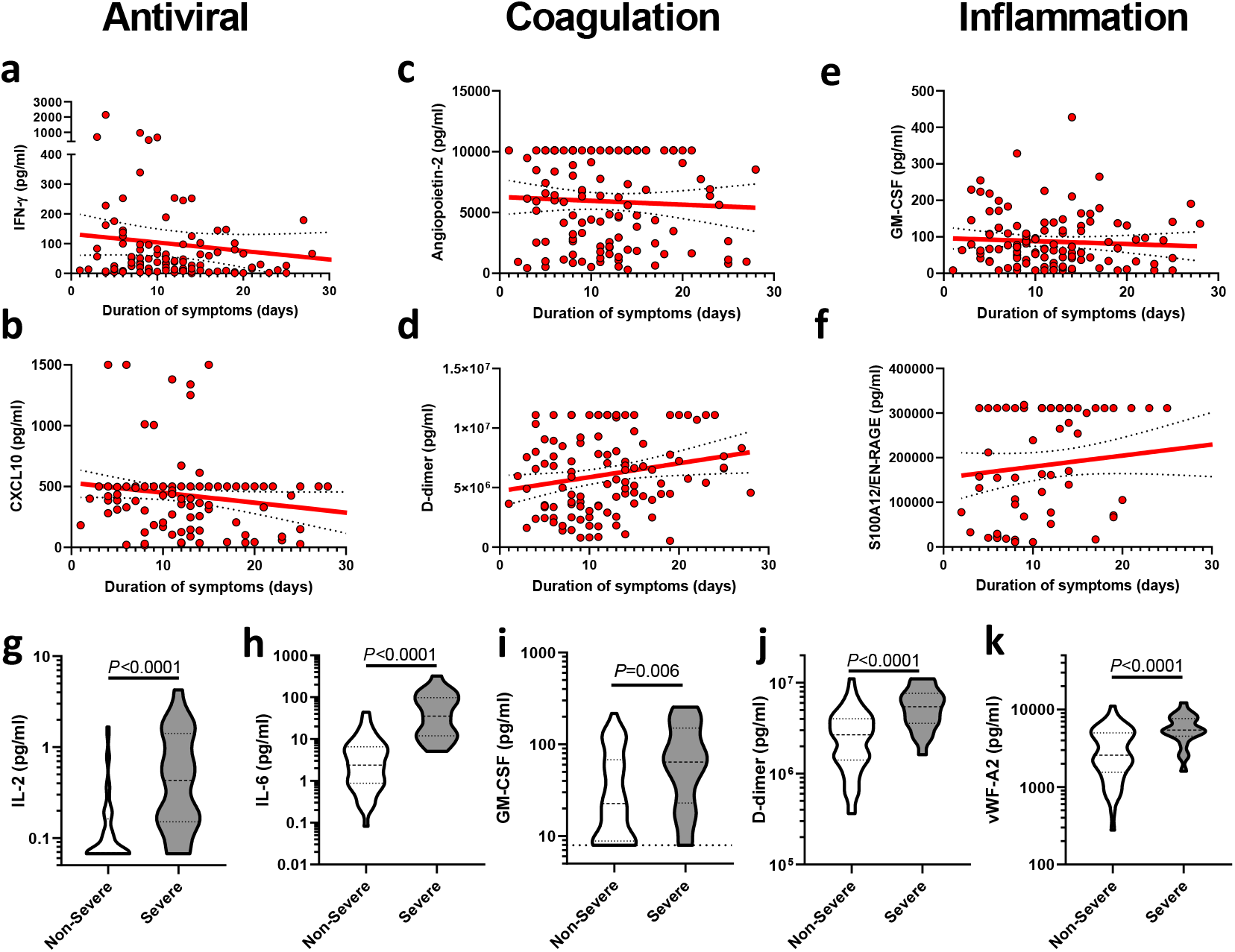
Longitudinal analysis of plasma mediator levels demonstrate a progressive immune response and an exaggerated signature of endothelial injury and inflammation early in fatal COVID-19. Plasma levels of a) IFN-γ, b) CXCL10, c) Angiopoietin-2, d) D-dimer, e) GM-CSF, and f) EN-RAGE/S100A12 over the course of disease in patients with fatal COVID-19. Plasma mediator levels of g) IL-2, h) IL-6, i) GM-CSF, j) D-dimer, and k) von-Willebrand factor A2 (vWF-A2) within the first 4 days of symptom onset in patients in severity groups 6/7 or 8 (“Severe”, n=22) and groups 3, 4, or 5 (“Non-Severe”, n=54). Linear regressions with 95% confidence intervals are shown in panels a-f. Data in panels g-k were analyzed for statistical significance using Mann-Whitney U tests, where thick horizontal dashed lines denote the median values and thin horizontal dashed lines denote the interquartile ranges.

Finally, we hypothesized that differences in plasma mediator levels between patients with Severe (groups 6/7 and 8) and Non-severe (groups 3, 4, and 5) COVID-19 would be apparent within the first few days of symptoms. Indeed, within the first 4 days of symptoms several mediators were significantly elevated in the Severe group, relative to Non-severe, including IL-2, IL-6, and GM-CSF (all *P*<0.0001, Fig. 4g-i, respectively), indicating a pronounced inflammatory response early in Severe disease. Similarly, many markers of coagulation and endothelial injury were elevated in Severe, relative to Non-severe, including D-dimer and vWF-A2 (*P*<0.0001, Fig. 4j and 4k, respectively), in addition to angiopoietin-2 and IL-1α (which can be activated by thrombin ^33^) (Supplementary Fig. 5). By comparison the lung damage-associated marker EN-RAGE ^30^ was not significantly different between the Severe and Non-severe groups in the first 4 days of symptoms (*P*=0.098, Supplementary Fig. 5). Together, these data indicated that severe COVID-19 is associated with elevated levels of plasma mediators indicative of coagulation, endothelial activation and a broad inflammatory response including CXCL10, GM-CSF, and IL-6. These differences were apparent within the first days of symptoms, while markers of lung damage may only become elevated later in disease, potentially indicating a pathological role for these processes and a window of opportunity for early immunomodulation to prevent significant lung damage.

While markers of fibrinolysis have previously been associated with disease severity ^16^ and thrombosis is common in severe and fatal COVID-19 ^4, 5, 20^ the causes of this manifestation of severe disease are not known. We demonstrate that increasing disease severity is associated with broad elevations in inflammatory mediator levels, alongside a signature of endothelial injury. This signal was most pronounced in fatal COVID-19 and was apparent even in the early stages of disease.

The elevation of angiopoietin-2, thrombomodulin, and vWF-A2 in fatal COVID-19 cases provides evidence for the involvement of endothelial injury in COVID-19 severity. Endothelial injury following inflammatory damage, including the increasingly recognized pulmonary artery vasculitis ^4, 20^ in COVID-19, may result in the initiation of a pro-coagulant role for these cells ^34^. Alternatively, this response could be triggered by direct viral infection of vascular cells (though this has yet to be conclusively determined ^34^ viral replication in non-respiratory tissues is commonly observed at post-mortem ^4, 7^); or thrombin mediated activation of IL-1α ^33^. This pro-coagulant role could lead to the deposition of microthrombi, evident in COVID-19 ^4^, the development of features of disseminated intravascular coagulopathy (DIC) and ultimately elevated levels D-dimer through the degradation of fibrin rich thrombi ^28^. Neutrophilic inflammation could have an etiological role in endothelial injury though neutrophilia is predominantly a feature of the later phases of COVID-19 ^1^, while endothelial injury was evident in the first days of symptoms. However, the continued thrombosis in late stage fatal COVID-19 may result from neutrophil mediated coagulation, observed in other settings ^35, 36, 37^ and recently demonstrated in COVID-19 ^38^. Combined, these results indicate a multiplicity of possible pro-coagulant triggers that may contribute to pathology at different stages of disease.

We found that the antiviral immune mediator CXCL10 and the myeloid cell growth factor GM-CSF, were strikingly elevated in fatal cases of COVID-19. This is confirmed by a recent report describing the potential utility of CXCL10 as an early prognostic marker of COVID-19 severity ^39^. An influx of monocytes/macrophages has been described in the lung parenchyma in fatal COVID-19, combined with a mononuclear cell pulmonary artery vasculitis ^6^, and presence of pro-inflammatory monocyte-derived macrophages in bronchoalveolar lavage fluid from patients with severe COVID-19 ^4, 40^. The elevation of CXCL10 and GM-CSF in severe disease reported here could contribute to monocyte recruitment and activation leading to this vasculitis, alongside the role of GM-CSF in the recruitment of neutrophils to the pulmonary vasculature ^41^.

Large scale randomized clinical trials for IL-6 signaling antagonists are on-going, though early results of the COVACTA trial of Tocilizumab found no improvement in clinical status or mortality ^42^. Small scale studies of anti-GM-CSF have shown promising results ^43, 44^ but require formal testing in a clinical trial. Given the role of GM-CSF in granulopoiesis and enhancement of neutrophil survival, alongside the neutrophil activation observed in late stage fatal COVID-19, these trials may inform our understanding of the importance of this pathway in COVID-19 immunopathogenesis ^45^. While early studies demonstrated elevated GM-CSF levels in both ICU and non-ICU treated COVID-19 patients ^1^, we now demonstrate a positive association with disease severity and outcome, in agreement with reports of elevated frequencies of GM-CSF^+^ Th1 cells in patients with COVID-19 requiring ICU treatment ^46^.

While many cytokines and other inflammatory mediators were most significantly elevated in fatal and critical COVID-19, these data do not necessarily support the concept of a “cytokine storm” in COVID-19 ^12, 13^. While some elements, such as elevated IL-6 and ferritin levels (reported in other studies, but not seen here) ^8, 9, 10^, are reminiscent of sHLH, the relatively gradual clinical progression and persistent elevation of some cytokines, even during the early stages of symptomatic disease, are uncommon amongst conditions associated with cytokine storms such as toxic-shock syndrome and bacterial sepsis.

To our knowledge, this is to date the largest study of inflammatory responses in COVID-19. The multicenter nature of ISARIC4C adds to the ability to interpret and apply these results to other settings. However, further studies are needed to determine the prognostic value of these key plasma biomarkers, including multivariable analyses of biological data alongside clinical and demographic data. This detailed level of analysis may also enable the phenotyping of patients most likely to respond to individual therapies. Future analyses should focus on the biological features of patients that respond to therapeutic interventions, such as dexamethasone ^2, 3^, to enable mechanistic insight and targeting of treatment. The clear distinction between patients that would progress to severe COVID-19 and those that would not, even in the earliest stages of disease, indicates that early therapeutic intervention may be crucial to limit mortality. Overall, these data indicate an early inflammatory response in COVID-19, most prominent in those who will later suffer severe or fatal disease. These responses may enable the development of prognostic biomarkers, inform our understanding of immunopathogenesis in COVID-19 and enable novel approaches for therapeutic intervention.

## Supplementary Methods

### Study design and setting

The ISARIC WHO Clinical Characterization Protocol for Severe Emerging Infections in the UK (CCP-UK) is an ongoing prospective cohort study of hospitalized patients with COVID-19, which is recruiting in 258 hospitals in England, Scotland, and Wales (National Institute for Health Research Clinical Research Network Central Portfolio Management System ID: 14152) ^47^. The protocol, revision history, case report form, patient information leaflets, consent forms and details of the Independent Data and Material Access Committee are available online ^48^ and published previously ^22^.

### Participants

Hospitalized patients with PCR-proven or high likelihood of SARS-CoV-2 infection were recruited, including both patients with community- and hospital-acquired COVID-19. This study analyzed plasma from blood samples obtained on the day of enrolment to the study (day 1, Tier 1) and additional serial samples obtained following a sampling schedule (Tier 2) harmonized with international investigators to allow meaningful comparison of results between studies ^21^. Healthy controls were recruited prior to December 2019 under approval from the London – Fulham Research Ethics Committee (REC) (reference 14/LO/1023) or from healthy donors following informed consent from a sub-collection of the Imperial College Healthcare NHS Trust National Institute for Health Research Imperial Biomedical Research Centre Tissue Bank. Use of the sub-collection was approved by the Tissue Bank Ethics Committee (Approval R12023). Samples from community managed COVID-19 cases were collected through a subproject of Imperial College London Communicable Disease Research Tissue Bank, under approval from the south central Oxford REC (reference 15/SC/0089).

### HScores

To calculate partial HScores ^24^, ferritin, triglyceride and AST measurements from this study were combined with recorded results from case report forms for temperature and routine hemoglobin, white cell counts, and platelet counts.

### Immunoassays

IFN-γ, TNF-α, IL-1β, IL-2, IL-4, IL-6, CXCL8/IL-8, IL-10, IL-12p70 and IL-13 were quantified using MSD (Mesoscale Diagnostics, Rockville, Maryland, USA) V-Plex proinflammatory plates on a SQ120 Quickplex instrument. IL-1α, IL-1ra, IL-6Rα, angiopoetin-1, angiopoetin-2, endothelin-1, VEGF-D, D-dimer, thrombomodulin, Tie2, von-Willebrand Factor-A2 (vWF-A2), G-CSF, GM-CSF, IL-17A, LCN2/NGAL, CXCL10/IP-10, CCL2, CCL3, CCL4 and CCL5 were quantified using a Bio Plex 200 instrument (Bio-Rad, Hercules, California, USA) with custom Luminex panel kits from Biotechne (Minneapolis, Minnesota, USA) and MilliporeSigma (Burlington, Massachusetts, USA). IFN-α was quantified using Quanterix (Billerica, Massachusetts, USA) IFN-α assay kits on the SIMOA platform. All values at or below the lower limit of detection (LLOD) were replaced with the geometric mean of the lower limits of detection across plates for each assay.

### Statistical analyses

Statistical analyses used GraphPad Prism v8.3.0 (GraphPad, La Jolla, California, USA) R version 3.6.1 and Python 3.7.3 with Pandas 1.0.3 and Seaborn 0.10.0. Non-parametric mediator data (as determined by D’Agostino and Pearson normality test) were analyzed by ANOVA using Kruskal-Wallis tests with Dunn’s test for multiple comparisons of patient groups within in time group. Non-parametric two-way analyses were performed using Mann-Whitney U tests. Correlation matrix analysis was performed using the R packages ggplot2 and ggcorrplot and Spearman’s test for correlation of non-parametric data, after *P*-value adjustment for multiple testing. The false discovery rate, or expected proportion of discoveries which are falsely rejected, was controlled using the methods of Benjamini and Hochberg. Heatmaps of scaled plasma mediator data were generated using the ComplexHeatmap package in R with rows and columns split by K-means clustering and dendrograms based on Ward’s minimum variance method (ward.D2) and Spearmans rank correlations. For heatmap analyses missing values were imputed by predictive mean matching using the Multivariate Imputation by Chained Equations (MICE) package ^49^.

## Data Availability

ISARIC4C data are available for request through an independent data and materials access committee. All other data are available upon reasonable request from the authors.

---

**Supplementary table 1.**
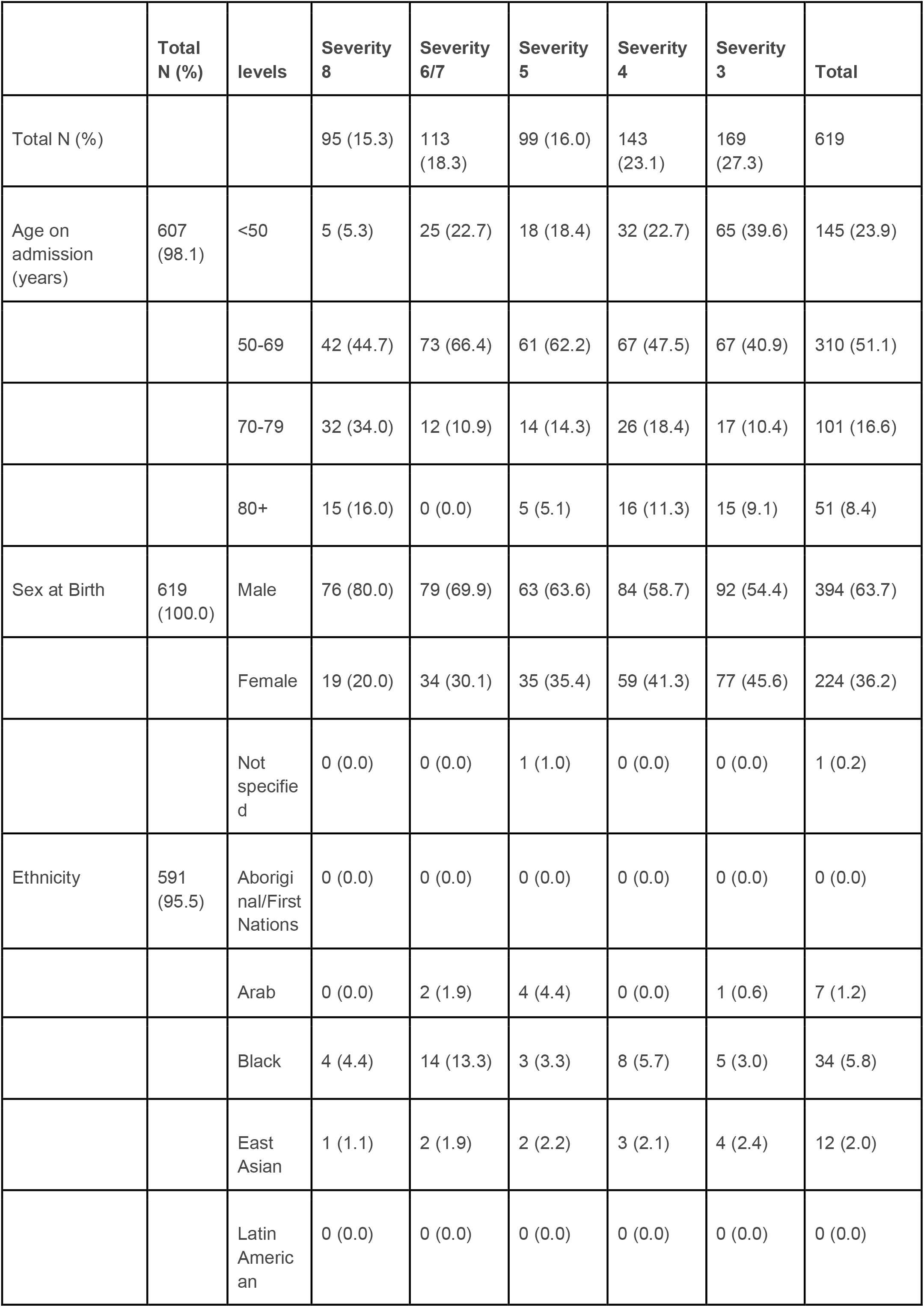
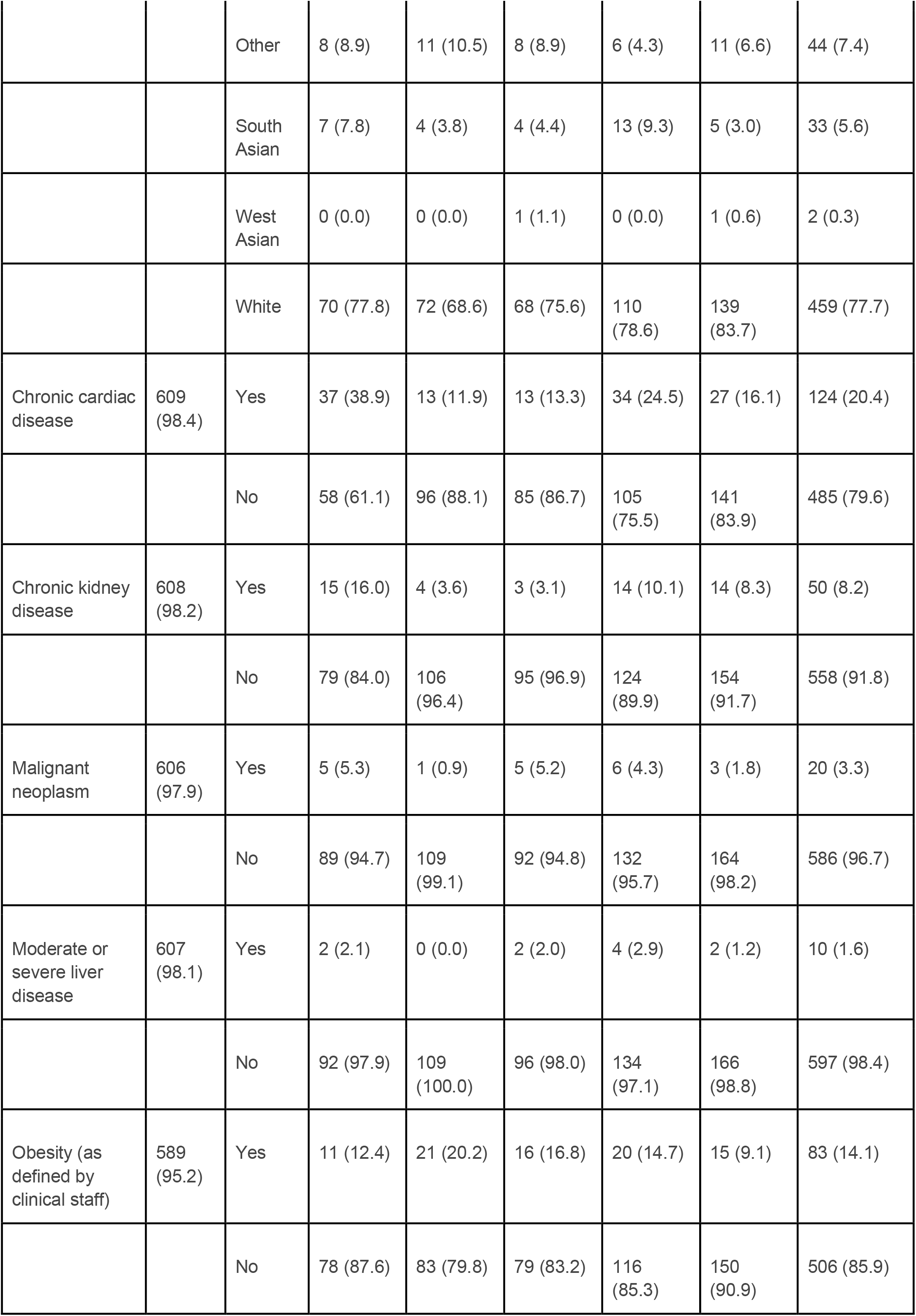
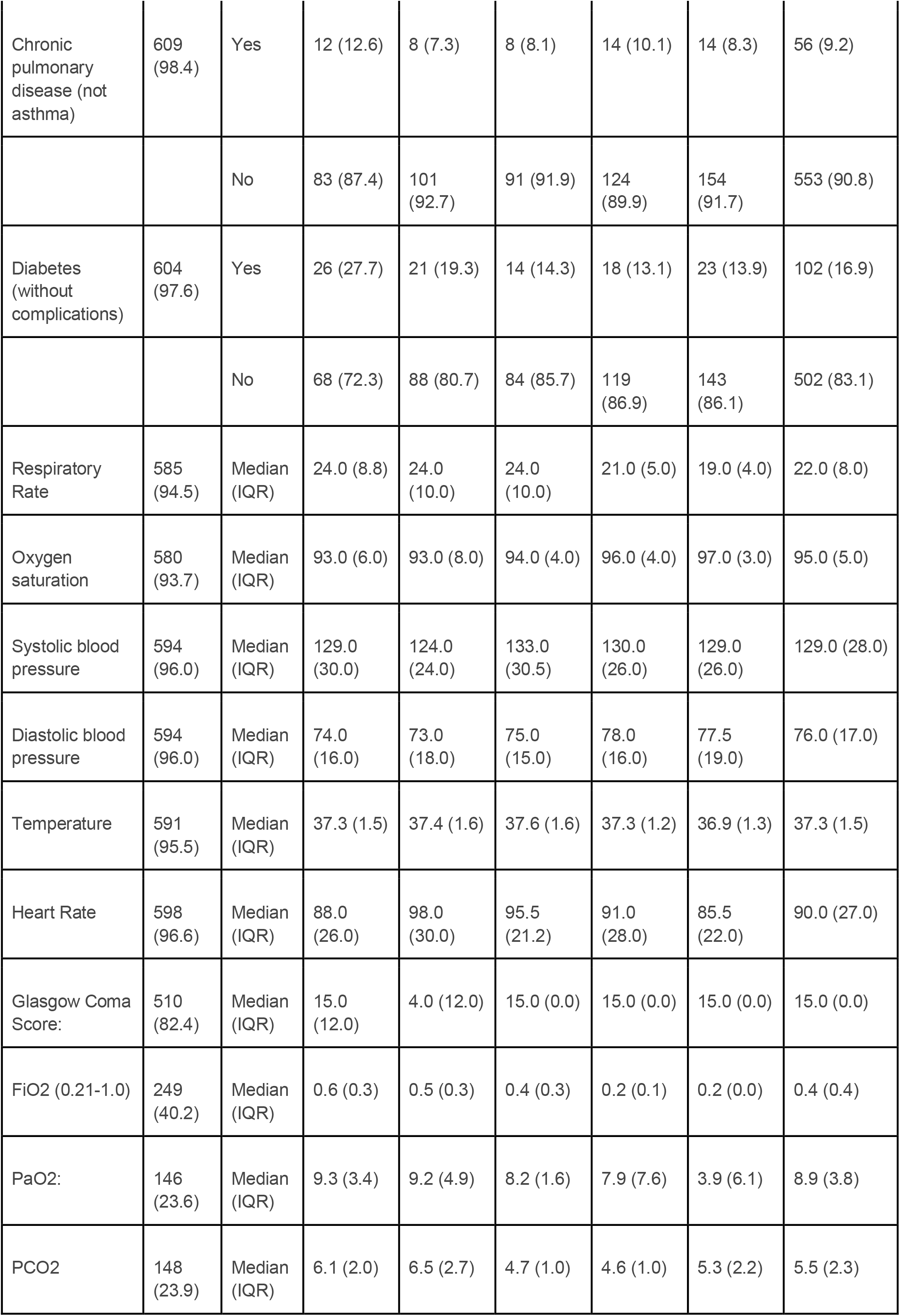
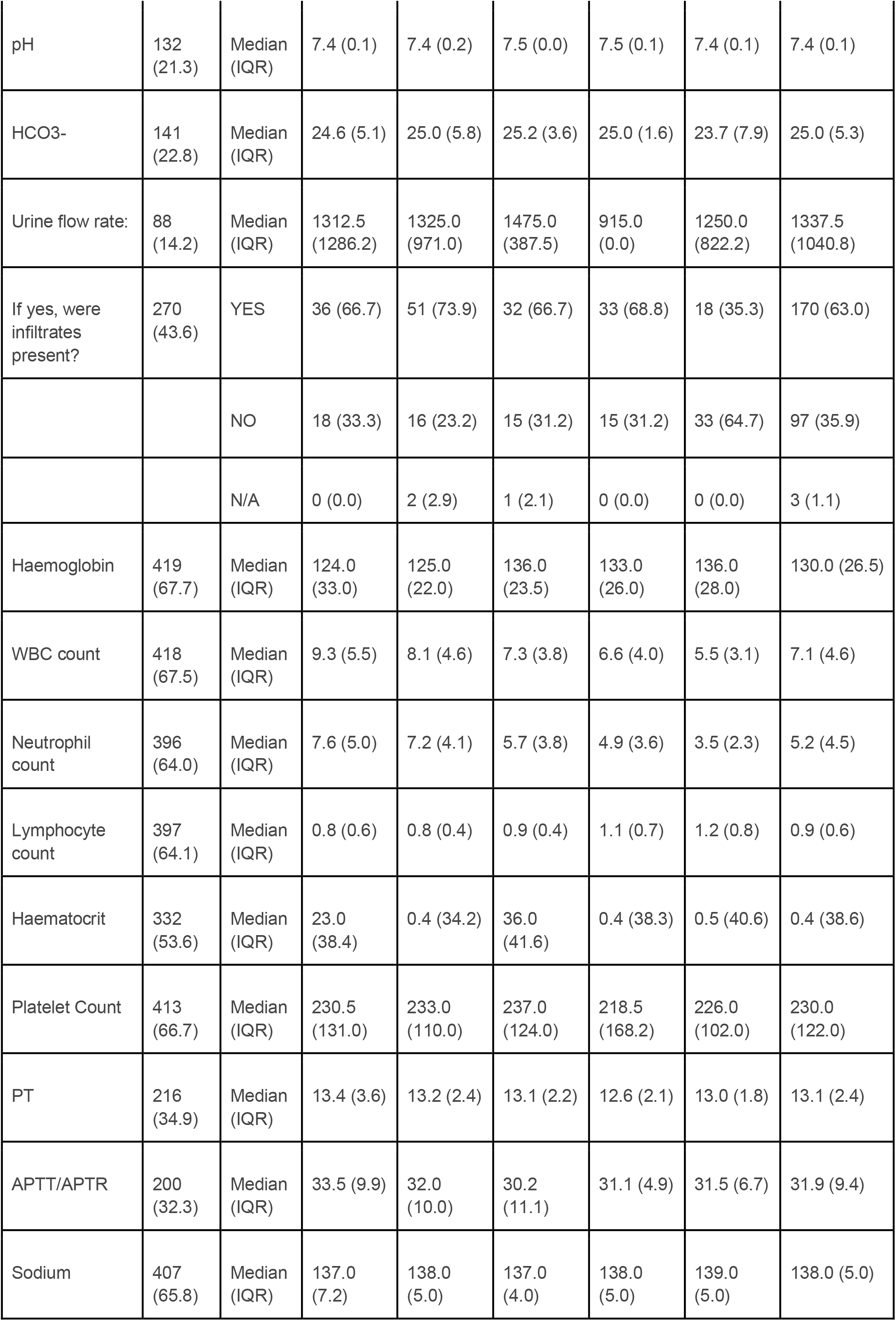
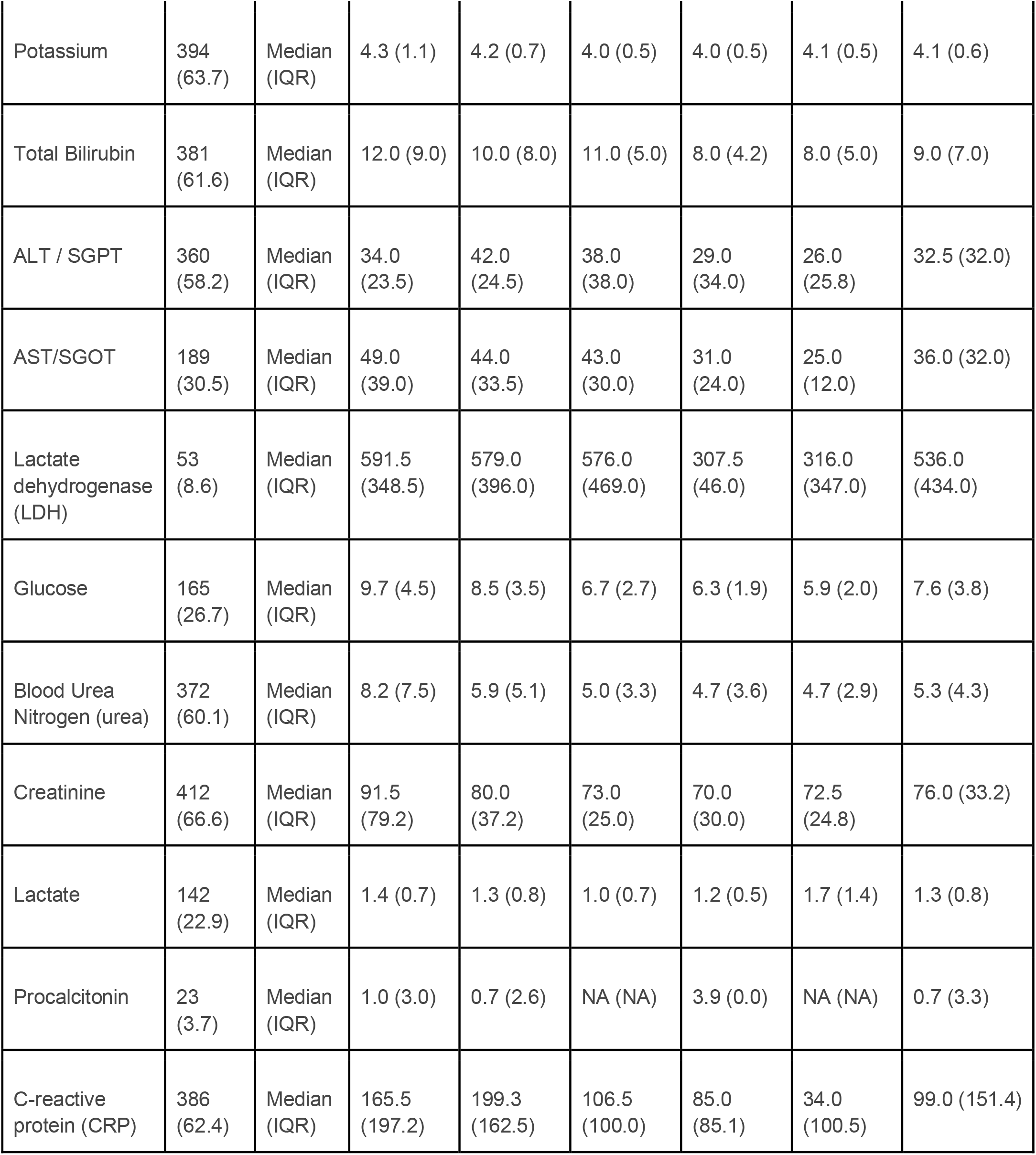
Clinical demographics, hematology, and biochemistry data of patients hospitalized with COVID-19 at the time of study enrolment.

|  | **Total**<br>**N (%)** | **levels** | **Severity**<br>**8** | **Severity**<br>**6/7** | **Severity**<br>**5** | **Severity**<br>**4** | **Severity**<br>**3** | **Total** |
| --- | --- | --- | --- | --- | --- | --- | --- | --- |
| Total N (%) |  |  | 95 (15.3) | 113<br>(18.3) | 99 (16.0) | 143<br>(23.1) | 169<br>(27.3) | 619 |
| Age on admission (years) | 607<br>(98.1) | <50 | 5 (5.3) | 25 (22.7) | 18 (18.4) | 32 (22.7) | 65 (39.6) | 145 (23.9) |
|  |  | 50-69 | 42 (44.7) | 73 (66.4) | 61 (62.2) | 67 (47.5) | 67 (40.9) | 310 (51.1) |
|  |  | 70-79 | 32 (34.0) | 12 (10.9) | 14 (14.3) | 26 (18.4) | 17 (10.4) | 101 (16.6) |
|  |  | 80+ | 15 (16.0) | 0 (0.0) | 5 (5.1) | 16 (11.3) | 15 (9.1) | 51 (8.4) |
| Sex at Birth | 619<br>(100.0) | Male | 76 (80.0) | 79 (69.9) | 63 (63.6) | 84 (58.7) | 92 (54.4) | 394 (63.7) |
|  |  | Female | 19 (20.0) | 34 (30.1) | 35 (35.4) | 59 (41.3) | 77 (45.6) | 224 (36.2) |
|  |  | Not specified | 0 (0.0) | 0 (0.0) | 1 (1.0) | 0 (0.0) | 0 (0.0) | 1 (0.2) |
| Ethnicity | 591<br>(95.5) | Aboriginal/First Nations | 0 (0.0) | 0 (0.0) | 0 (0.0) | 0 (0.0) | 0 (0.0) | 0 (0.0) |
|  |  | Arab | 0 (0.0) | 2 (1.9) | 4 (4.4) | 0 (0.0) | 1 (0.6) | 7 (1.2) |
|  |  | Black | 4 (4.4) | 14 (13.3) | 3 (3.3) | 8 (5.7) | 5 (3.0) | 34 (5.8) |
|  |  | East Asian | 1 (1.1) | 2 (1.9) | 2 (2.2) | 3 (2.1) | 4 (2.4) | 12 (2.0) |
|  |  | Latin American | 0 (0.0) | 0 (0.0) | 0 (0.0) | 0 (0.0) | 0 (0.0) | 0 (0.0) |
|  |  | Other | 8 (8.9) | 11 (10.5) | 8 (8.9) | 6 (4.3) | 11 (6.6) | 44 (7.4) |
|  |  | South Asian | 7 (7.8) | 4 (3.8) | 4 (4.4) | 13 (9.3) | 5 (3.0) | 33 (5.6) |
|  |  | West Asian | 0 (0.0) | 0 (0.0) | 1 (1.1) | 0 (0.0) | 1 (0.6) | 2 (0.3) |
|  |  | White | 70 (77.8) | 72 (68.6) | 68 (75.6) | 110 (78.6) | 139 (83.7) | 459 (77.7) |
| Chronic cardiac disease | 609 (98.4) | Yes | 37 (38.9) | 13 (11.9) | 13 (13.3) | 34 (24.5) | 27 (16.1) | 124 (20.4) |
|  |  | No | 58 (61.1) | 96 (88.1) | 85 (86.7) | 105 (75.5) | 141 (83.9) | 485 (79.6) |
| Chronic kidney disease | 608 (98.2) | Yes | 15 (16.0) | 4 (3.6) | 3 (3.1) | 14 (10.1) | 14 (8.3) | 50 (8.2) |
|  |  | No | 79 (84.0) | 106 (96.4) | 95 (96.9) | 124 (89.9) | 154 (91.7) | 558 (91.8) |
| Malignant neoplasm | 606 (97.9) | Yes | 5 (5.3) | 1 (0.9) | 5 (5.2) | 6 (4.3) | 3 (1.8) | 20 (3.3) |
|  |  | No | 89 (94.7) | 109 (99.1) | 92 (94.8) | 132 (95.7) | 164 (98.2) | 586 (96.7) |
| Moderate or severe liver disease | 607 (98.1) | Yes | 2 (2.1) | 0 (0.0) | 2 (2.0) | 4 (2.9) | 2 (1.2) | 10 (1.6) |
|  |  | No | 92 (97.9) | 109 (100.0) | 96 (98.0) | 134 (97.1) | 166 (98.8) | 597 (98.4) |
| Obesity (as defined by clinical staff) | 589 (95.2) | Yes | 11 (12.4) | 21 (20.2) | 16 (16.8) | 20 (14.7) | 15 (9.1) | 83 (14.1) |
|  |  | No | 78 (87.6) | 83 (79.8) | 79 (83.2) | 116 (85.3) | 150 (90.9) | 506 (85.9) |
| Chronic pulmonary disease (not asthma) | 609 (98.4) | Yes | 12 (12.6) | 8 (7.3) | 8 (8.1) | 14 (10.1) | 14 (8.3) | 56 (9.2) |
|  |  | No | 83 (87.4) | 101 (92.7) | 91 (91.9) | 124 (89.9) | 154 (91.7) | 553 (90.8) |
| Diabetes (without complications) | 604 (97.6) | Yes | 26 (27.7) | 21 (19.3) | 14 (14.3) | 18 (13.1) | 23 (13.9) | 102 (16.9) |
|  |  | No | 68 (72.3) | 88 (80.7) | 84 (85.7) | 119 (86.9) | 143 (86.1) | 502 (83.1) |
| Respiratory Rate | 585 (94.5) | Median (IQR) | 24.0 (8.8) | 24.0 (10.0) | 24.0 (10.0) | 21.0 (5.0) | 19.0 (4.0) | 22.0 (8.0) |
| Oxygen saturation | 580 (93.7) | Median (IQR) | 93.0 (6.0) | 93.0 (8.0) | 94.0 (4.0) | 96.0 (4.0) | 97.0 (3.0) | 95.0 (5.0) |
| Systolic blood pressure | 594 (96.0) | Median (IQR) | 129.0 (30.0) | 124.0 (24.0) | 133.0 (30.5) | 130.0 (26.0) | 129.0 (26.0) | 129.0 (28.0) |
| Diastolic blood pressure | 594 (96.0) | Median (IQR) | 74.0 (16.0) | 73.0 (18.0) | 75.0 (15.0) | 78.0 (16.0) | 77.5 (19.0) | 76.0 (17.0) |
| Temperature | 591 (95.5) | Median (IQR) | 37.3 (1.5) | 37.4 (1.6) | 37.6 (1.6) | 37.3 (1.2) | 36.9 (1.3) | 37.3 (1.5) |
| Heart Rate | 598 (96.6) | Median (IQR) | 88.0 (26.0) | 98.0 (30.0) | 95.5 (21.2) | 91.0 (28.0) | 85.5 (22.0) | 90.0 (27.0) |
| Glasgow Coma Score: | 510 (82.4) | Median (IQR) | 15.0 (12.0) | 4.0 (12.0) | 15.0 (0.0) | 15.0 (0.0) | 15.0 (0.0) | 15.0 (0.0) |
| FiO2 (0.21-1.0) | 249 (40.2) | Median (IQR) | 0.6 (0.3) | 0.5 (0.3) | 0.4 (0.3) | 0.2 (0.1) | 0.2 (0.0) | 0.4 (0.4) |
| PaO2: | 146 (23.6) | Median (IQR) | 9.3 (3.4) | 9.2 (4.9) | 8.2 (1.6) | 7.9 (7.6) | 3.9 (6.1) | 8.9 (3.8) |
| PCO2 | 148 (23.9) | Median (IQR) | 6.1 (2.0) | 6.5 (2.7) | 4.7 (1.0) | 4.6 (1.0) | 5.3 (2.2) | 5.5 (2.3) |
| pH | 132<br>(21.3) | Median<br>(IQR) | 7.4 (0.1) | 7.4 (0.2) | 7.5 (0.0) | 7.5 (0.1) | 7.4 (0.1) | 7.4 (0.1) |
| HCO<sub>3</sub><sup>-</sup> | 141<br>(22.8) | Median<br>(IQR) | 24.6 (5.1) | 25.0 (5.8) | 25.2 (3.6) | 25.0 (1.6) | 23.7 (7.9) | 25.0 (5.3) |
| Urine flow rate: | 88<br>(14.2) | Median<br>(IQR) | 1312.5<br>(1286.2) | 1325.0<br>(971.0) | 1475.0<br>(387.5) | 915.0<br>(0.0) | 1250.0<br>(822.2) | 1337.5<br>(1040.8) |
| If yes, were<br>infiltrates<br>present? | 270<br>(43.6) | YES | 36 (66.7) | 51 (73.9) | 32 (66.7) | 33 (68.8) | 18 (35.3) | 170 (63.0) |
|  |  | NO | 18 (33.3) | 16 (23.2) | 15 (31.2) | 15 (31.2) | 33 (64.7) | 97 (35.9) |
|  |  | N/A | 0 (0.0) | 2 (2.9) | 1 (2.1) | 0 (0.0) | 0 (0.0) | 3 (1.1) |
| Haemoglobin | 419<br>(67.7) | Median<br>(IQR) | 124.0<br>(33.0) | 125.0<br>(22.0) | 136.0<br>(23.5) | 133.0<br>(26.0) | 136.0<br>(28.0) | 130.0 (26.5) |
| WBC count | 418<br>(67.5) | Median<br>(IQR) | 9.3 (5.5) | 8.1 (4.6) | 7.3 (3.8) | 6.6 (4.0) | 5.5 (3.1) | 7.1 (4.6) |
| Neutrophil<br>count | 396<br>(64.0) | Median<br>(IQR) | 7.6 (5.0) | 7.2 (4.1) | 5.7 (3.8) | 4.9 (3.6) | 3.5 (2.3) | 5.2 (4.5) |
| Lymphocyte<br>count | 397<br>(64.1) | Median<br>(IQR) | 0.8 (0.6) | 0.8 (0.4) | 0.9 (0.4) | 1.1 (0.7) | 1.2 (0.8) | 0.9 (0.6) |
| Haematocrit | 332<br>(53.6) | Median<br>(IQR) | 23.0<br>(38.4) | 0.4 (34.2) | 36.0<br>(41.6) | 0.4 (38.3) | 0.5 (40.6) | 0.4 (38.6) |
| Platelet Count | 413<br>(66.7) | Median<br>(IQR) | 230.5<br>(131.0) | 233.0<br>(110.0) | 237.0<br>(124.0) | 218.5<br>(168.2) | 226.0<br>(102.0) | 230.0<br>(122.0) |
| PT | 216<br>(34.9) | Median<br>(IQR) | 13.4 (3.6) | 13.2 (2.4) | 13.1 (2.2) | 12.6 (2.1) | 13.0 (1.8) | 13.1 (2.4) |
| APTT/APTR | 200<br>(32.3) | Median<br>(IQR) | 33.5 (9.9) | 32.0<br>(10.0) | 30.2<br>(11.1) | 31.1 (4.9) | 31.5 (6.7) | 31.9 (9.4) |
| Sodium | 407<br>(65.8) | Median<br>(IQR) | 137.0<br>(7.2) | 138.0<br>(5.0) | 137.0<br>(4.0) | 138.0<br>(5.0) | 139.0<br>(5.0) | 138.0 (5.0) |
| Potassium | 394<br>(63.7) | Median<br>(IQR) | 4.3 (1.1) | 4.2 (0.7) | 4.0 (0.5) | 4.0 (0.5) | 4.1 (0.5) | 4.1 (0.6) |
| Total Bilirubin | 381<br>(61.6) | Median<br>(IQR) | 12.0 (9.0) | 10.0 (8.0) | 11.0 (5.0) | 8.0 (4.2) | 8.0 (5.0) | 9.0 (7.0) |
| ALT / SGPT | 360<br>(58.2) | Median<br>(IQR) | 34.0<br>(23.5) | 42.0<br>(24.5) | 38.0<br>(38.0) | 29.0<br>(34.0) | 26.0<br>(25.8) | 32.5 (32.0) |
| AST/SGOT | 189<br>(30.5) | Median<br>(IQR) | 49.0<br>(39.0) | 44.0<br>(33.5) | 43.0<br>(30.0) | 31.0<br>(24.0) | 25.0<br>(12.0) | 36.0 (32.0) |
| Lactate<br>dehydrogenase<br>(LDH) | 53<br>(8.6) | Median<br>(IQR) | 591.5<br>(348.5) | 579.0<br>(396.0) | 576.0<br>(469.0) | 307.5<br>(46.0) | 316.0<br>(347.0) | 536.0<br>(434.0) |
| Glucose | 165<br>(26.7) | Median<br>(IQR) | 9.7 (4.5) | 8.5 (3.5) | 6.7 (2.7) | 6.3 (1.9) | 5.9 (2.0) | 7.6 (3.8) |
| Blood Urea<br>Nitrogen (urea) | 372<br>(60.1) | Median<br>(IQR) | 8.2 (7.5) | 5.9 (5.1) | 5.0 (3.3) | 4.7 (3.6) | 4.7 (2.9) | 5.3 (4.3) |
| Creatinine | 412<br>(66.6) | Median<br>(IQR) | 91.5<br>(79.2) | 80.0<br>(37.2) | 73.0<br>(25.0) | 70.0<br>(30.0) | 72.5<br>(24.8) | 76.0 (33.2) |
| Lactate | 142<br>(22.9) | Median<br>(IQR) | 1.4 (0.7) | 1.3 (0.8) | 1.0 (0.7) | 1.2 (0.5) | 1.7 (1.4) | 1.3 (0.8) |
| Procalcitonin | 23<br>(3.7) | Median<br>(IQR) | 1.0 (3.0) | 0.7 (2.6) | NA (NA) | 3.9 (0.0) | NA (NA) | 0.7 (3.3) |
| C-reactive<br>protein (CRP) | 386<br>(62.4) | Median<br>(IQR) | 165.5<br>(197.2) | 199.3<br>(162.5) | 106.5<br>(100.0) | 85.0<br>(85.1) | 34.0<br>(100.5) | 99.0 (151.4) |

**Supplementary Fig. 1.**
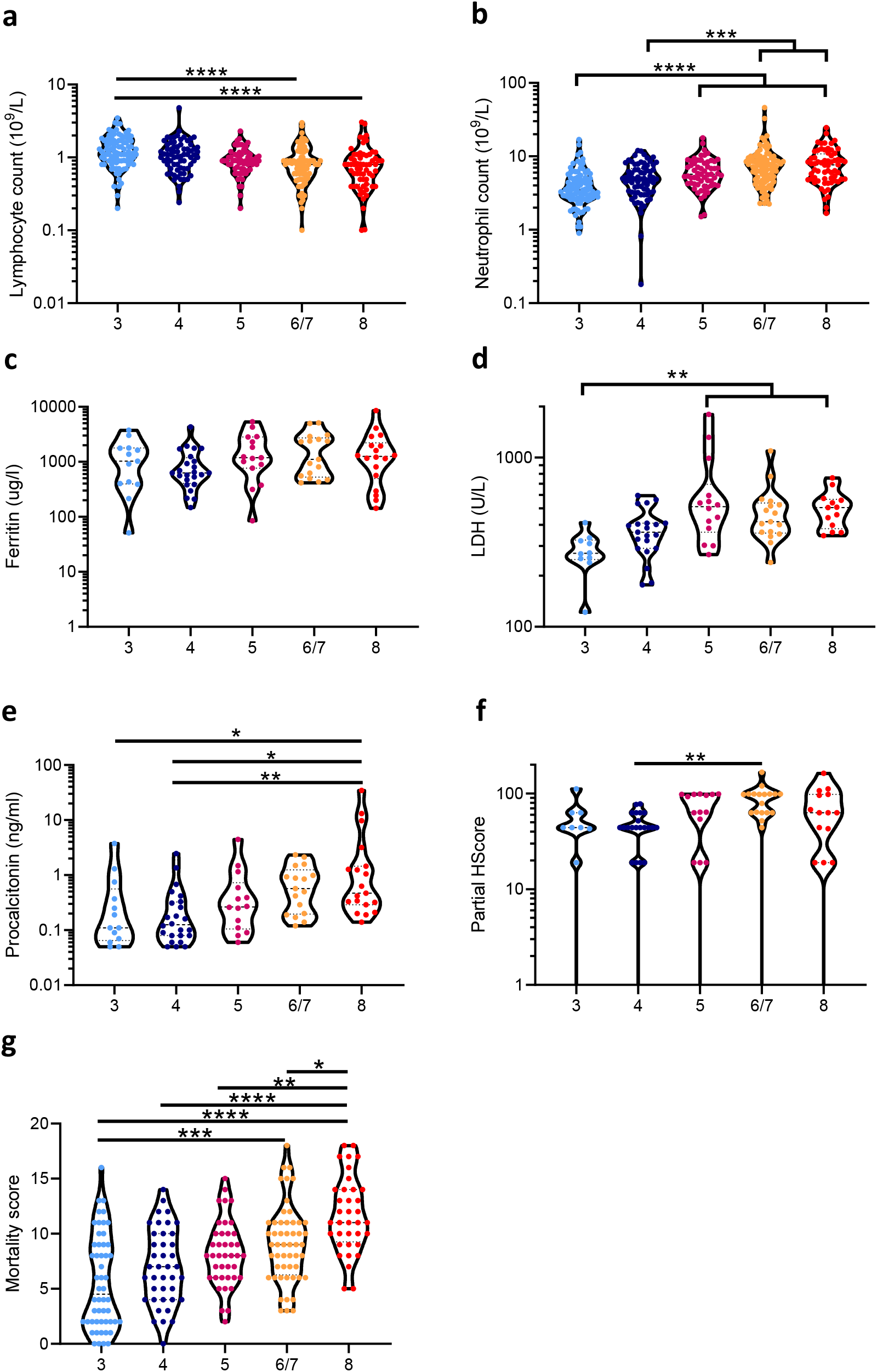
Clinical hematology, biochemistry, and severity scores of patients hospitalized with COVID-19 at enrolment. a) Peripheral blood lymphocyte count, b) neutrophil count, c) ferritin levels, d) lactate dehydrogenase (LDH) levels, e) procalcitonin levels, f) partial HScores, and g) ISARIC4C mortality scores at the time of enrolment in hospitalized patients with COVID-19 that would: not require oxygen support (‘3’, n=9-93); require an oxygen face mask (‘4’, n=22-71); require non-invasive ventilation or high-flow nasal cannulae (‘5’, n=15-63); require invasive mechanical ventilation (‘6/7’, n=19-91); or progress to fatal disease (‘8’, n=15-63). Data were analyzed for statistical significance using Kruskal-Wallis tests with Dunn’s tests for multiple comparisons between all groups. \**P*<0.05, \*\**P*<0.01, \*\*\**P*<0.001, \*\*\*\**P*<0.0001.

**Supplementary Fig. 2.**
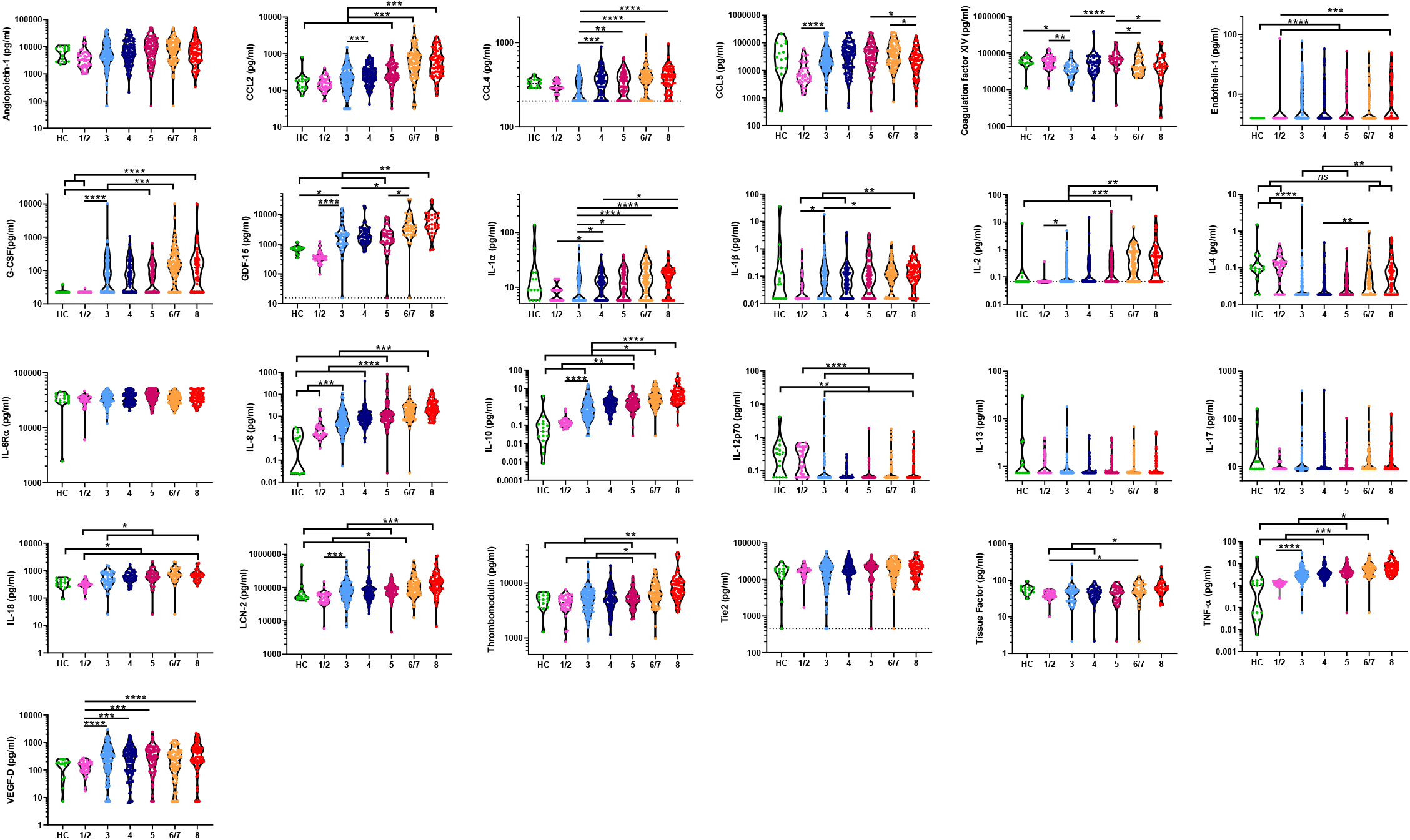
Plasma mediators at the time of enrolment in patients hospitalized with COVID-19. Mediator levels were quantified from plasma collected at the point of study enrolment from hospitalized patients with COVID-19 that would: not require oxygen support (‘3’, n=128), require an oxygen face mask (‘4’, n=103), require non-invasive ventilation or high-flow nasal cannulae (‘5’, n=78), require invasive mechanical ventilation (‘6/7’, n=87) or progress to fatal disease (‘8’, n=69). Data were analyzed for statistical significance using Kruskal-Wallis tests with Dunn’s tests for multiple comparisons between all groups. \**P*<0.05, \*\**P*<0.01, \*\*\**P*<0.001, \*\*\*\**P*<0.0001.

**Supplementary Fig. 3.**
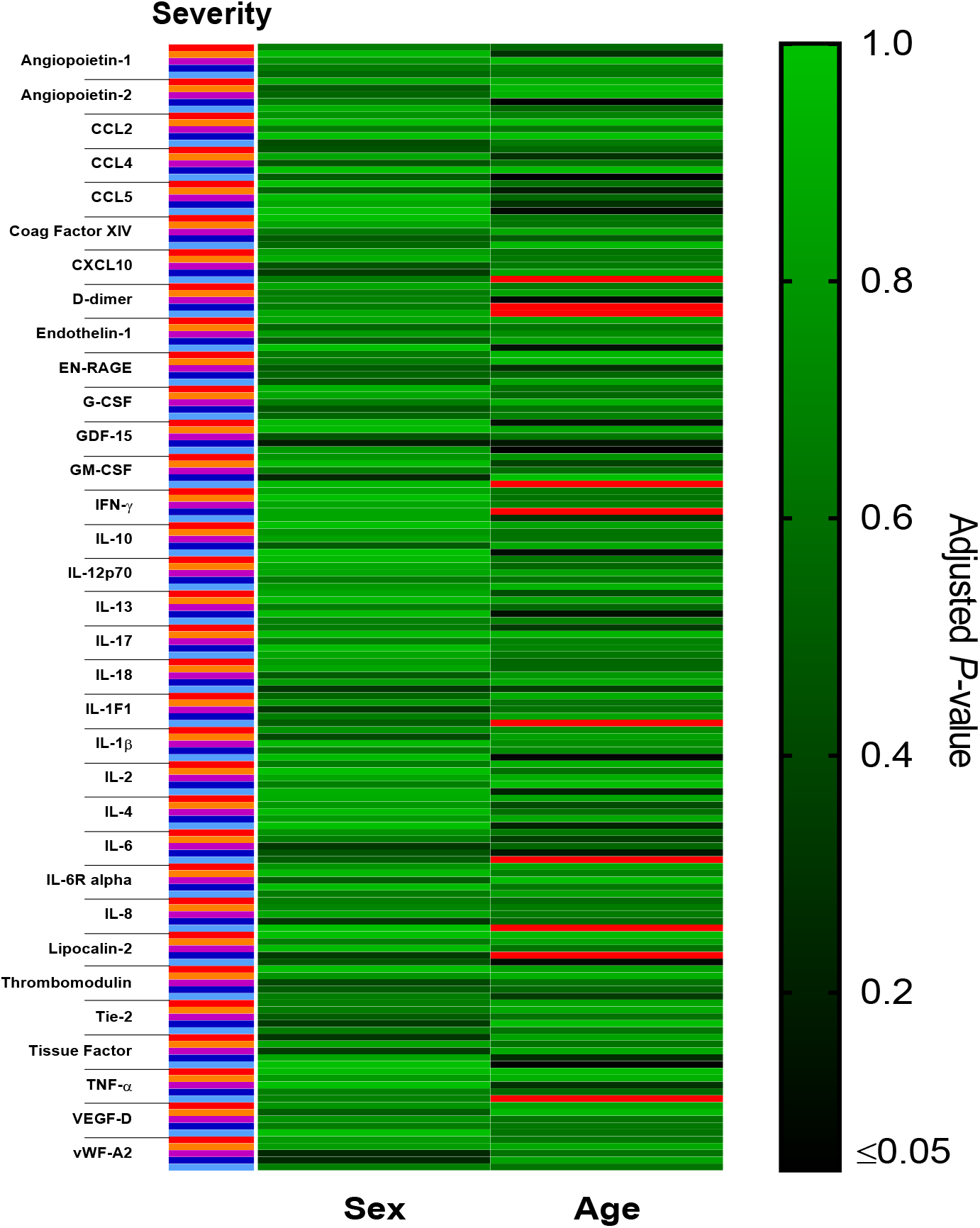
Age, but not sex, is associated with differences in plasma cytokine levels within COVID-19 disease outcome groups. Heatmap of false-discovery rate adjusted *P*-values for each plasma mediator between males and females (“Sex”) and those aged ≥70 years and <70 years (“Age”) within each disease outcome group (‘8’=Red, ‘6/7’=Orange, ‘5’=Purple, ‘4’=Dark blue, ‘3’=Cyan). Data were analyzed using Mann-Whitney U tests with *P*-value adjustment for false discovery rate.

**Supplementary Fig. 4.**
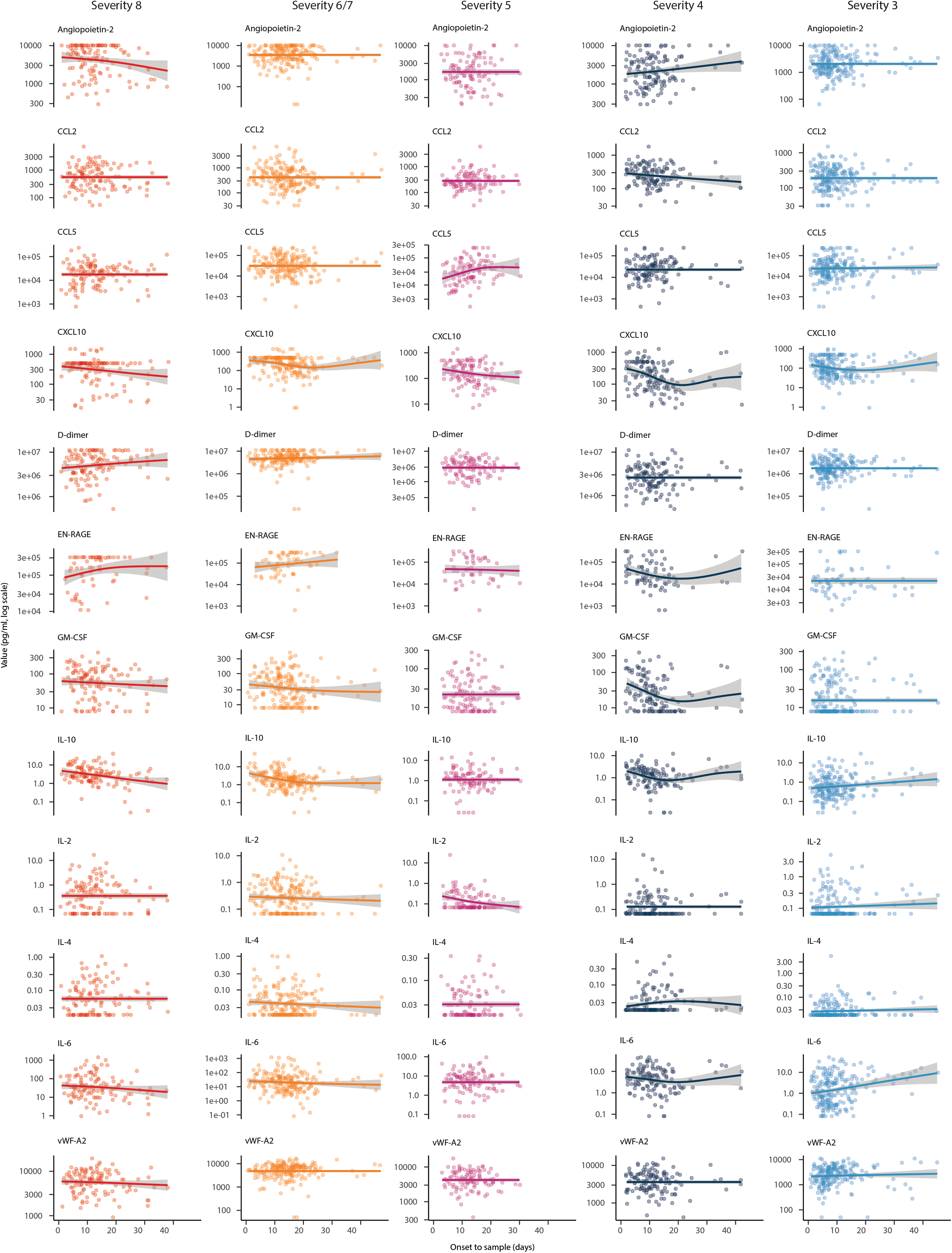
Longitudinal analysis of selected plasma mediators within each disease outcome group. All data within each severity group was related to the duration of symptoms at the time of sample collection (“Onset to sample”, measured in days) for each plasma mediator. Generalized additive modelling was used to fit a restricted cubic spline which is plotted together with the standard error (grey).

**Supplementary Fig. 5.**
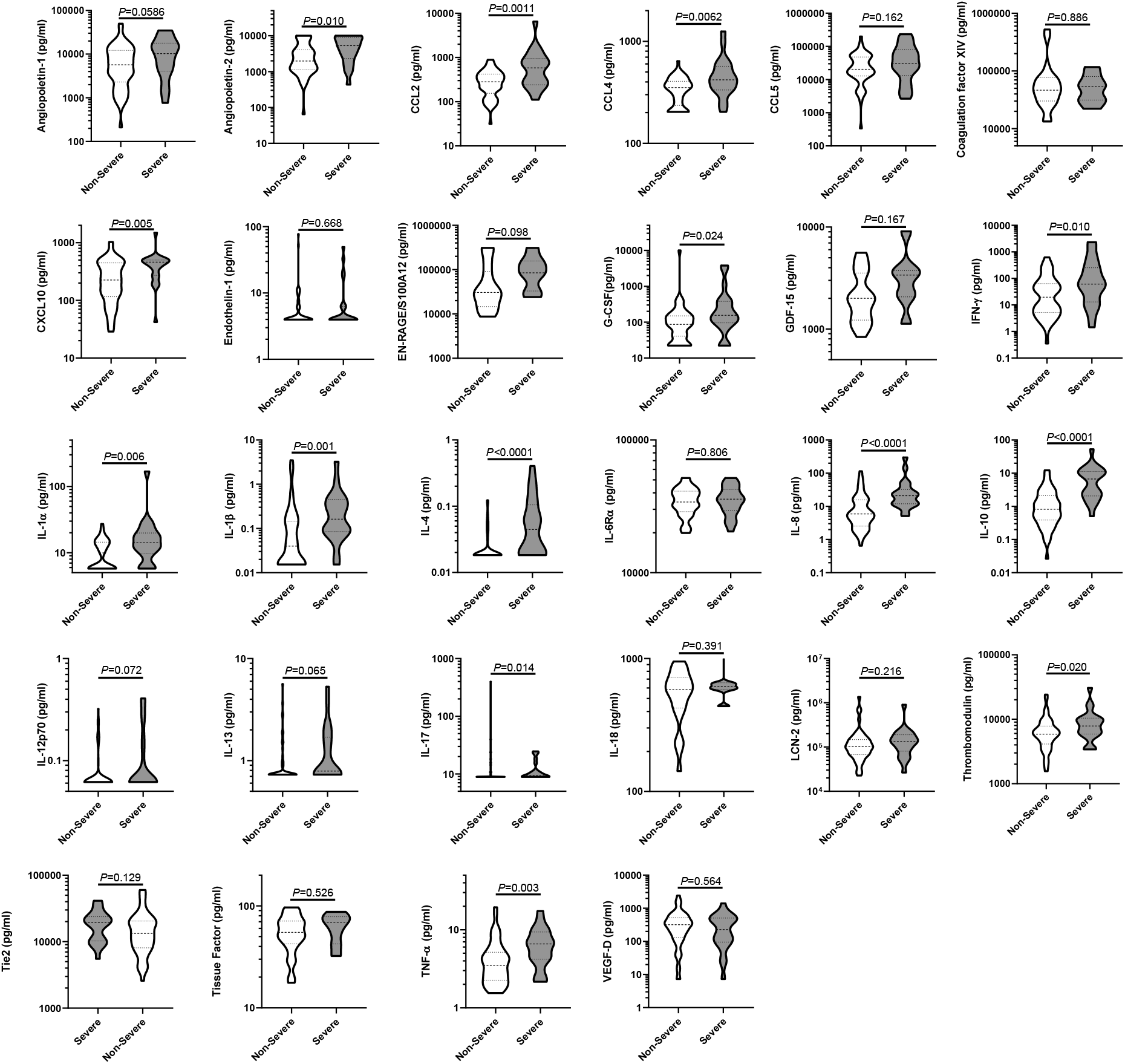
Longitudinal analysis of selected plasma mediators within each disease outcome group. Levels of immune mediators collected within the first 4 days of symptom onset in patients in the groups 6/7 or 8 (“Severe”, n=22) and groups 3, 4, or 5 (“Non-Severe”, n=54). Data were analyzed for statistical significance using Mann-Whitney U tests, where thick horizontal dashed lines denote the median values and thin horizontal dashed lines denote the interquartile ranges.

## The ISARIC4C Investigators

### Consortium Lead Investigator

J Kenneth Baillie, *Chief Investigator:* Malcolm G Semple, *Co-Lead Investigator:* Peter JM Openshaw. *ISARIC Clinical Coordinator:* Gail Carson.

### Co-Investigators

Beatrice Alex, Benjamin Bach, Wendy S Barclay, Debby Bogaert, Meera Chand, Graham S Cooke, Annemarie B Docherty, Jake Dunning, Ana da Silva Filipe, Tom Fletcher, Christopher A Green, Ewen M Harrison, Julian A Hiscox, Antonia Ying Wai Ho, Peter W Horby, Samreen Ijaz, Saye Khoo, Paul Klenerman, Andrew Law, Wei Shen Lim, Alexander J Mentzer, Laura Merson, Alison M Meynert, Mahdad Noursadeghi, Shona C Moore, Massimo Palmarini, William A Paxton, Georgios Pollakis, Nicholas Price, Andrew Rambaut, David L Robertson, Clark D Russell, Vanessa Sancho-Shimizu, Janet T Scott, Thushan de Silva, Louise Sigfrid, Tom Solomon, Shiranee Sriskandan, David Stuart, Charlotte Summers, Richard S Tedder, Emma C Thomson, AA Roger Thompson, Ryan S Thwaites, Lance CW Turtle, Maria Zambon.

### Project Managers

Hayley Hardwick, Chloe Donohue, Ruth Lyons, Fiona Griffiths, Wilna Oosthuyzen.

### Data Analysts

Lisa Norman, Riinu Pius, Tom M Drake, Cameron J Fairfield, Stephen Knight, Kenneth A Mclean, Derek Murphy, Catherine A Shaw.

### Data and Information System Manager

Jo Dalton, Michelle Girvan, Egle Saviciute, Stephanie Roberts, Janet Harrison, Laura Marsh, Marie Connor, Sophie Halpin, Clare Jackson, Carrol Gamble.

### Data integration and presentation

Gary Leeming, Andrew Law, Murray Wham, Sara Clohisey, Ross Hendry, James Scott-Brown.

### Material Management

William Greenhalf, Victoria Shaw, Sarah McDonald.

### Patient engagement

Seán Keating

### Outbreak Laboratory Staff and Volunteers

Katie A. Ahmed, Jane A Armstrong, Milton Ashworth, Innocent G Asiimwe, Siddharth Bakshi, Samantha L Barlow, Laura Booth, Benjamin Brennan, Katie Bullock, Benjamin WA Catterall, Jordan J Clark, Emily A Clarke, Sarah Cole, Louise Cooper, Helen Cox, Christopher Davis, Oslem Dincarslan, Chris Dunn, Philip Dyer, Angela Elliott, Anthony Evans, Lorna Finch, Lewis WS Fisher, Terry Foster, Isabel Garcia-Dorival, Willliam Greenhalf, Philip Gunning, Catherine Hartley, Antonia Ho, Rebecca L Jensen, Christopher B Jones, Trevor R Jones, Shadia Khandaker, Katharine King, Robyn T. Kiy, Chrysa Koukorava, Annette Lake, Suzannah Lant, Diane Latawiec, L Lavelle-Langham, Daniella Lefteri, Lauren Lett, Lucia A Livoti, Maria Mancini, Sarah McDonald, Laurence McEvoy, John McLauchlan, Soeren Metelmann, Nahida S Miah, Joanna Middleton, Joyce Mitchell, Shona C Moore, Ellen G Murphy, Rebekah Penrice-Randal, Jack Pilgrim, Tessa Prince, Will Reynolds, P. Matthew Ridley, Debby Sales, Victoria E Shaw, Rebecca K Shears, Benjamin Small, Krishanthi S Subramaniam, Agnieska Szemiel, Aislynn Taggart, Jolanta Tanianis-Hughes, Jordan Thomas, Erwan Trochu, Libby van Tonder, Eve Wilcock, J. Eunice Zhang.

### Local Principal Investigators

Kayode Adeniji, Daniel Agranoff, Ken Agwuh, Dhiraj Ail, Ana Alegria, Brian Angus, Abdul Ashish, Dougal Atkinson, Shahedal Bari, Gavin Barlow, Stella Barnass, Nicholas Barrett, Christopher Bassford, David Baxter, Michael Beadsworth, Jolanta Bernatoniene, John Berridge, Nicola Best, Pieter Bothma, David Brealey, Robin Brittain-Long, Naomi Bulteel, Tom Burden, Andrew Burtenshaw, Vikki Caruth, David Chadwick, Duncan Chambler, Nigel Chee, Jenny Child, Srikanth Chukkambotla, Tom Clark, Paul Collini, Catherine Cosgrove, Jason Cupitt, Maria-Teresa Cutino-Moguel, Paul Dark, Chris Dawson, Samir Dervisevic, Phil Donnison, Sam Douthwaite, Ingrid DuRand, Ahilanadan Dushianthan, Tristan Dyer, Cariad Evans, Chi Eziefula, Chrisopher Fegan, Adam Finn, Duncan Fullerton, Sanjeev Garg, Sanjeev Garg, Atul Garg, Effrossyni Gkrania-Klotsas, Jo Godden, Arthur Goldsmith, Clive Graham, Elaine Hardy, Stuart Hartshorn, Daniel Harvey, Peter Havalda, Daniel B Hawcutt, Maria Hobrok, Luke Hodgson, Anita Holme, Anil Hormis, Michael Jacobs, Susan Jain, Paul Jennings, Agilan Kaliappan, Vidya Kasipandian, Stephen Kegg, Michael Kelsey, Jason Kendall, Caroline Kerrison, Ian Kerslake, Oliver Koch, Gouri Koduri, George Koshy, Shondipon Laha, Steven Laird, Susan Larkin, Tamas Leiner, Patrick Lillie, James Limb, Vanessa Linnett, Jeff Little, Michael MacMahon, Emily MacNaughton, Ravish Mankregod, Huw Masson, Elijah Matovu, Katherine McCullough, Ruth McEwen, Manjula Meda, Gary Mills, Jane Minton, Mariyam Mirfenderesky, Kavya Mohandas, Quen Mok, James Moon, Elinoor Moore, Patrick Morgan, Craig Morris, Katherine Mortimore, Samuel Moses, Mbiye Mpenge, Rohinton Mulla, Michael Murphy, Megan Nagel, Thapas Nagarajan, Mark Nelson, Igor Otahal, Mark Pais, Selva Panchatsharam, Hassan Paraiso, Brij Patel, Justin Pepperell, Mark Peters, Mandeep Phull, Stefania Pintus, Jagtur Singh Pooni, Frank Post, David Price, Rachel Prout, Nikolas Rae, Henrik Reschreiter, Tim Reynolds, Neil Richardson, Mark Roberts, Devender Roberts, Alistair Rose, Guy Rousseau, Brendan Ryan, Taranprit Saluja, Aarti Shah, Prad Shanmuga, Anil Sharma, Anna Shawcross, Jeremy Sizer, Manu Shankar-Hari, Richard Smith, Catherine Snelson, Nick Spittle, Nikki Staines, Tom Stambach, Richard Stewart, Pradeep Subudhi, Tamas Szakmany, Kate Tatham, Jo Thomas, Chris Thompson, Robert Thompson, Ascanio Tridente, Darell Tupper-Carey, Mary Twagira, Andrew Ustianowski, Nick Vallotton, Lisa Vincent-Smith, Shico Visuvanathan, Alan Vuylsteke, Sam Waddy, Rachel Wake, Andrew Walden, Ingeborg Welters, Tony Whitehouse, Paul Whittaker, Ashley Whittington, Meme Wijesinghe, Martin Williams, Lawrence Wilson, Sarah Wilson, Stephen Winchester, Martin Wiselka, Adam Wolverson, Daniel G Wooton, Andrew Workman, Bryan Yates, and Peter Young.

## Acknowledgements

This work uses data provided by patients and collected by the NHS as part of their care and support #DataSavesLives. We are extremely grateful to the 2,648 frontline NHS clinical and research staff and volunteer medical students, who collected this data in challenging circumstances; and the generosity of the participants and their families for their individual contributions in these difficult times.

We also acknowledge the support of Jeremy J Farrar, Nahoko Shindo, Devika Dixit, Nipunie Rajapakse, Lyndsey Castle, Martha Buckley, Debbie Malden, Katherine Newell, Kwame O’Neill, Emmanuelle Denis, Claire Petersen, Scott Mullaney, Sue MacFarlane, Nicole Maziere, Julien Martinez, Oslem Dincarslan, Annette Lake, Lucy Cook, Robert Hammond, Rachael Quinlan, Benjamin Harris, Mohamed Zuhair, Michael Fertleman, Claudia Selck, Danai Koftori, Alex Cocker, Zainab Saeed, Isaac Day-Weber, Aris Aristodemou, Patricia Watber, and Yunchuan Ding.

